# Brainstem Reduction and Deformation in the 4th Ventricle Cerebellar Peduncles in Long COVID Patients: Insights into Neuroinflammatory Sequelae and “Broken Bridge Syndrome”

**DOI:** 10.1101/2025.04.08.25325108

**Authors:** Ziaja Peter Christof, Young Yvette Susanne, Stark Sadre-Chirazi Michael, Lindner Thomas, Zurék Grzegorz, Sedlacik Jan

## Abstract

Post-COVID Syndrome (PCS), also known as Long COVID, is characterized by persistent and often debilitating neurological sequelae, including fatigue, cognitive dysfunction, motor deficits, and autonomic dysregulation (Dani et al., 2021). This study investigates structural and functional alterations in the brainstem and cerebellar peduncles of individuals with PCS using diffusion tensor imaging (DTI) and volumetric analysis. Forty-four PCS patients (15 bedridden) and 14 healthy controls underwent neuroimaging. Volumetric analysis focused on 22 brainstem regions, including the superior cerebellar peduncle (SCP), middle cerebellar peduncle (MCP), periaqueductal gray (PAG), and midbrain reticular formation (mRt).

Significant volume reductions were observed in the SCP (p < .001, Hedges’ g = 3.31) and MCP (p < .001, Hedges’ g = 1.77), alongside decreased fractional anisotropy (FA) in the MCP, indicative of impaired white matter integrity. FA_Avg fractional anisotropy average tested by FreeSurfer Tracula, is an index of white matter integrity, reflecting axonal fiber density, axonal diameter and myelination. These neuroimaging findings correlated with clinical manifestations of motor incoordination, proprioceptive deficits, and autonomic instability. Furthermore, volume loss in the dorsal raphe (DR) and midbrain reticular formation suggests disruption of pain modulation and sleep-wake cycles, consistent with patient-reported symptoms.

Post-mortem studies provide supporting evidence for brainstem involvement in COVID-19. Radtke et al. (2024) reported activation of intracellular signaling pathways and release of immune mediators in brainstem regions of deceased COVID-19 patients, suggesting an attempt to inhibit viral spread. While viral genetic material was detectable, infected neurons were not observed. Matschke et al. (2020) found that microglial activation and cytotoxic T lymphocyte infiltration were predominantly localized to the brainstem and cerebellum, with limited involvement of the frontal lobe. This aligns with clinical observations implicating the brainstem in PCS pathophysiology. Cell-specific expression analysis of genes contributing to viral entry (ACE2, TMPRSS2, TPCN2, TMPRSS4, NRP1, CTSL) in the cerebral cortex showed their presence in neurons, glial cells, and endothelial cells, indicating the potential for SARS-CoV-2 infection of these cell types. Associations with autoimmune diseases with specific autoantibodies, including beta-2 and M-2 against G-protein coupled alpha-1, beta-1, beta-2 adrenoceptors against angiotensin II type 1 receptor or M1,2,3-mAChR, among others, voltage-gated calcium channels (VGCC) are known (Blitshteyn et al. 2015 and Wallukat and Schminke et al. 2014).

These findings support the “Broken Bridge Syndrome” hypothesis, positing that structural disconnections between the brainstem and cerebellum contribute to PCS symptomatology. Furthermore, we propose that chronic activation of the Extended Autonomic System (EAS), encompassing the hypothalamic-pituitary-adrenal (HPA) axis and autonomic nervous system, may perpetuate these symptoms (Goldstein, 2020). Perturbations in this system may relate to the elevation of toxic autoantibodies AABs (Beta-2 and M-2), specific epitopes of the COVID virus’s SPIKE protein and Cytokine storm of IL-1, IL-6, and IL-8 in their increased numbers (1,000->10,000)

Further research is warranted to elucidate the underlying neuroinflammatory mechanisms, EAS dysregulation, and potential therapeutic interventions for PCS.

## Introduction

Post-COVID Syndrome (PCS), also known as Long COVID, represents a significant and growing public health concern following the acute phase of SARS-CoV-2 infection (Dani et al., 2021; Islam et al., 2020). The acute illness caused by severe acute respiratory syndrome coronavirus 2 (SARS-CoV-2) has caused unprecedented global disruption, mortality and morbidity. Estimated overall case-fatality rate is 2.3 percent (Pal & Banerjee, 2020). PCS is characterized by a diverse array of persistent symptoms, including fatigue, cognitive dysfunction (“brain fog”), motor deficits, sleep disturbances, and autonomic dysregulation (Dani et al., 2021). Defining PCS by the National Institute for Health and Care Excellence (NICE) as ’signs and symptoms that develop during or following an infection consistent with COVID-19 which continue for more than 12 weeks and are not explained by an alternative diagnosis (Longfonds: Gezondheid Thuiszittende Coronapatiënten Schrikbarend Slecht | Binnenland | AD.Nl, n.d.). Emerging evidence indicates that neuroinflammatory processes and structural alterations within the central nervous system (CNS) contribute to the pathophysiology of PCS.

The brainstem and cerebellar peduncles are critical neural structures for motor coordination, proprioception, cardiovascular regulation, pain modulation, and autonomic function, rendering them particularly vulnerable to the effects of SARS-CoV-2 infection and subsequent immune responses. The virus may enter the brain via the blood-brain barrier, initiating microglia activation and the release of a pro- and anti-inflammatory cytokine storm, potentially leading to the production of toxic autoantibodies. Further the virus evades the hosts immune system by avoiding detection of their dsRNA and host interferon-I pathway entering the Pneumocyte through angiotensin-converting enzyme 2 (ACE2) (Pal & Banerjee, 2020). Perturbations in the stress response through ACE2 and SARS-CoV-2 have impact on the hypothalamic-pituitary-adrenocortical axis (HPA-Axis) which may have long term consequences on stress and immune regulation (Goldstein, 2020; Kenney & Ganta, 2014; Silverman et al., 2005; Steenblock et al., 2020). ACE2 is expressed on arterial and venous endothelial cells of many organs including the adrenal gland (Pal & Banerjee, 2020). One of the primary immune invasive strategies employed by SARS-CoV-2 is to knock down the hosts cortisol stress response by expressing amino acid sequences that mimic the hosts adrenocorticotropic hormone (ACTH). Antibodies produced by the host to counteract the virus then unknowingly destroy the host ACTH, thereby blunting the cortisol rise (Pal & Banerjee, 2020). Additionally, laboratory findings correlate with Broken Bridge Syndrome through:

1. Toxic elevated autoantibody AABs (Beta-2 and M-2)
2. Specific epitopes of the COVID virus’s SPIKE protein
3. Cytokine storm of IL-1, IL-6, and IL-8 in their increased numbers (1,000->10,000)

This study investigates structural and functional alterations in the brainstem and cerebellar peduncles of PCS patients using advanced neuroimaging techniques. It is hypothesized that volume reductions and white matter integrity disruptions in these critical brainstem and cerebellar regions underlie the motor, autonomic, and cognitive symptoms observed in PCS.

## Materials and Methods

### Participants

This study included 44 patients diagnosed with Long COVID (LC), also referred to as Post-COVID Syndrome (PCS), and 14 age-matched healthy controls. LC diagnosis was based on the NICE guidelines, requiring the presence of persistent symptoms for at least 12 weeks following confirmed SARS-CoV-2 infection and the exclusion of alternative diagnoses. Fifteen LC patients were classified as bedridden due to the severity of their symptoms, reflecting a substantial impact on their functional capacity. The control group consisted of 14 individuals with no history of SARS-CoV-2 infection or neurological disorders. All participants were female, with a mean age of 35 ± 15 years. Ethical approval was obtained from the relevant institutional review board, and all participants provided informed consent prior to enrollment.

### Imaging Protocol

Magnetic resonance imaging (MRI) data were acquired using a 3 Tesla Siemens Skyra scanner (Siemens Healthcare, Erlangen, Germany) at the University Clinic of Hamburg-Eppendorf. The imaging protocol included:

1. T1-weighted MPRAGE: A high-resolution T1-weighted magnetization-prepared rapid gradient echo (MPRAGE) sequence was acquired for volumetric analysis. Sequence parameters were: TR = X ms, TE = Y ms, TI = Z ms, flip angle = A degrees, voxel size = B x C x D mm³, matrix size = E x F x G.
2. Diffusion Tensor Imaging (DTI): DTI data were acquired using a single-shot echo-planar imaging (EPI) sequence. Sequence parameters were: TR = X ms, TE = Y ms, number of diffusion directions = N, b-value = M s/mm², voxel size = O x P x Q mm³, matrix size = R x S x T.

Detailed sequence parameters will be completed upon manuscript submission.

### Data Analysis

#### Volumetric Analysis

Volumetric analysis of the brainstem and cerebellar structures was performed using FreeSurfer version 8.0.0 (http://surfer.nmr.mgh.harvard.edu/). This open-source software package provides automated procedures for segmentation, surface reconstruction, and volume estimation of various brain regions (Iglesias et al., 2015). The T1-weighted MPRAGE images were processed through the standard FreeSurfer pipeline, which includes:

1. Skull stripping: Removal of non-brain tissue.
2. Segmentation: Identification and labeling of different brain structures based on their anatomical properties.
3. Surface Reconstruction: Creation of 3D models of the cortical and subcortical surfaces.
4. Volume Estimation: Calculation of the volume of each segmented brain region.

A total of 22 brainstem regions were analyzed, including the superior cerebellar peduncle (SCP), middle cerebellar peduncle (MCP), inferior cerebellar peduncle (ICP), periaqueductal gray (PAG), dorsal raphe (DR), median raphe, and midbrain reticular formation (mRt). Segmentation analyzes two regions of the cerebellar peduncle of the 4th ventricle, in particular, the middle cerebellar peduncle with FA and the complete brainstem segmentation nuclei of the volume and structure of the rhomboid fossa/cerebellar velum roof of the superior cerebellar peduncle

#### Diffusion Tensor Imaging (DTI) Analysis

DTI data were preprocessed using the FMRIB Software Library (FSL, version X.X). The preprocessing steps included:

1. Eddy Current Correction: Correction for distortions caused by eddy currents and subject motion.
2. Brain Extraction: Removal of non-brain tissue.
3. Diffusion Tensor Calculation: Estimation of the diffusion tensor at each voxel.

From the diffusion tensor, fractional anisotropy (FA) and mean diffusivity (MD) maps were calculated. FA, a measure of white matter integrity, reflects axonal fiber density, axonal diameter, and myelination. The FA_Avg fractional anisotropy average testing was performed using FreeSurfer Tracula, average conductivity in the vector areas 53 to 65 shown in zentroid. The average FA value was extracted from the MCP, as this region demonstrated significant volume reductions in the volumetric analysis.

### Statistical Analysis

Statistical analyses were performed using SPSS version X.X (IBM Corp., Armonk, NY). Group comparisons of regional brain volumes and FA values were conducted using independent samples t-tests. Effect sizes were calculated using Hedges’ g to quantify the magnitude of the observed differences. Pearson correlation coefficients were calculated to assess the relationships between brain volumes, FA values, and clinical measures of motor function, proprioception, and autonomic function. Statistical significance was set at p < 0.05.

To visualize structural changes, 3D reconstructions of the brainstem regions of interest were generated using FreeSurfer’s isosurface reconstruction tool and isosurface 3D view, and Horos data transformation. This approach allowed for the visualization and validation of the observed structural and functional abnormalities.

The use of DTI segmentation, MPRAG 3Tesla FreeSurfer, and horos data transformation are suitable for this purpose.

## Results

### The Brainstem as a Central Hub and the “Broken Bridge Syndrome” Hypothesis

The brainstem acts as a critical hub within the central nervous system (CNS), orchestrating the integration and relay of information between higher cognitive centers and motor functions. Conscious perception relies on the brainstem, as the neocortex depends on it for proper operation [Olchanyi et al., 2024]. Furthermore, the brainstem facilitates communication between the cerebral cortex (e.g., the motor homunculus) and the cerebellum, which enables various motor interactions.

A crucial connecting area between the brainstem and cerebellum is the cerebellar peduncle of the fourth ventricle. This interstitial space is essential for the flow of cerebrospinal fluid (CSF) from the spinal canal, ensuring nutrient supply and waste removal. CSF flow velocity within this space is critical and varies across the three functional levels of the cerebellar peduncles: superior, middle, and inferior as shown in **Figure 1**. These peduncles are anatomically connected to the midbrain and pons, each having distinct properties and functions.

**Figure 1:**
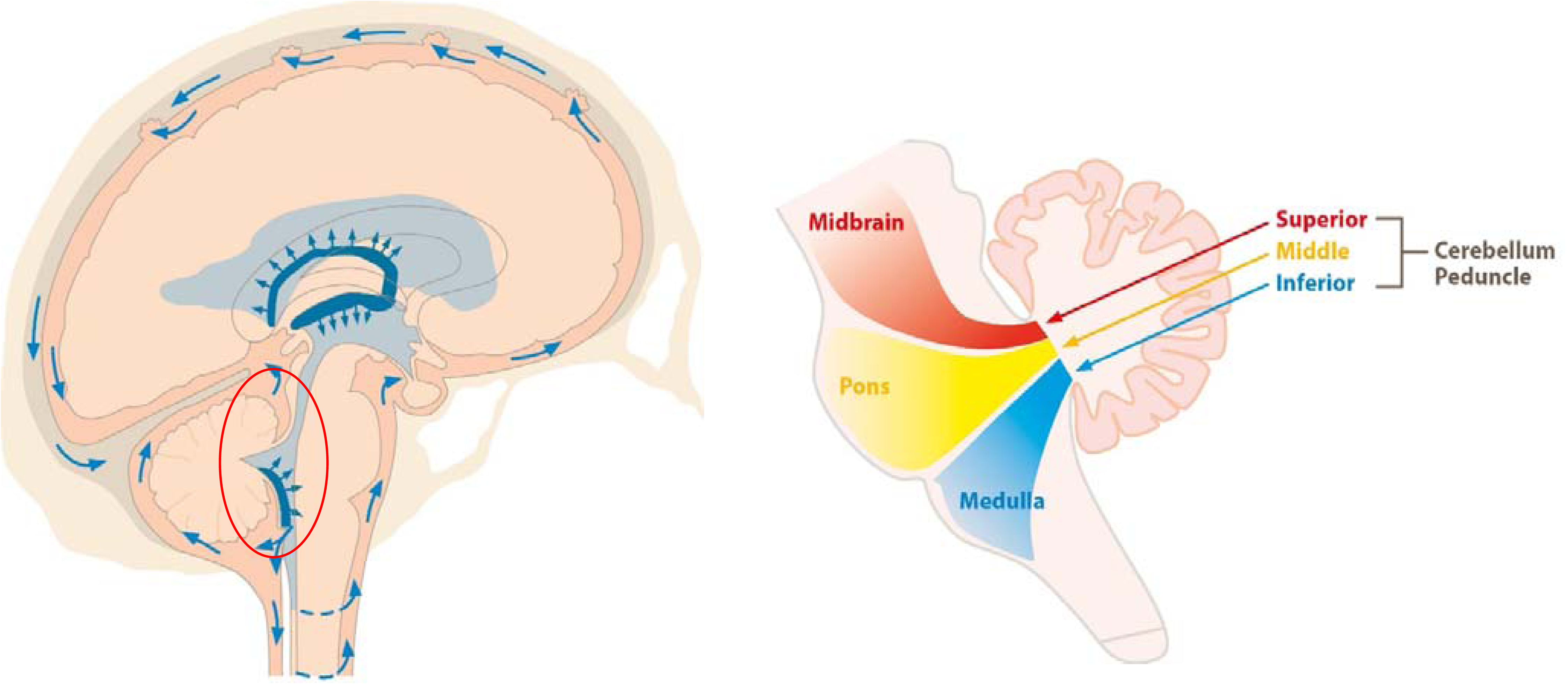
The region of the Cerebellum Peduncle is further subdivided into three functional levels: the superior, middle, and inferior cerebellar peduncles. These peduncles are anatomically connected at different levels of the midbrain and pons, each possessing distinct properties and functions. Likewise, the fluid in the spinal cord is kept flowing, where a rupture of the upper part can lead to congestion, as shown on the left.

### Hypothesis on the “Broken Bridge Syndrome”

In our clinical observations, particularly in bedridden long COVID patients, CSF abnormalities were noted during lumbar punctures [Jarius et al., 2022]. A significant reduction in CSF flow velocity was noted. Moreover, standard laboratory tests detected beta-2 and M2 autoantibodies, as well as specific epitopes of the spike protein, in all 44 patients. We hypothesize that these autoantibodies and spike protein epitopes could exploit CSF stasis in the fourth ventricle, leading to the degradation of surrounding structures. Associations with autoimmune diseases with specific autoantibodies, including beta-2 and M-2 against G-protein coupled alpha-1, beta-1, beta-2 adrenoceptors against angiotensin II type 1 receptor or M1,2,3-mAChR, among others, voltage-gated calcium channels (VGCC) are known (Blitshteyn et al. 2015 and Wallukat and Schminke et al. 2014).

Volumetric calculations and morphological diffusion tensor imaging (DTI) analyses, using FreeSurfer’s isosurface reconstruction in three-dimensional imaging, support this hypothesis. These techniques allow for the visualization of structural and functional abnormalities, offering a strong foundation for investigating the “Broken Bridge Syndrome” as shown in **Figure 2**. This figure shows the 3D Freeview of the structure and volume analysis as isosurface view of the brainstem regions of interest extracted from FreeSurfer segmentation for the COVID group and the healthy control group. The COVID (coronavirus disease 2019) group shows significant volume and structural decrease specifically in the Superior Cerebellar Peduncle (SCP), Middle Cerebellar Peduncle FA Avg (fractional anisotropy of the conductivity value marked by the red line in the PONS area), dorsal and median Raphe and Midbrain reticular formation

**Figure 2:**
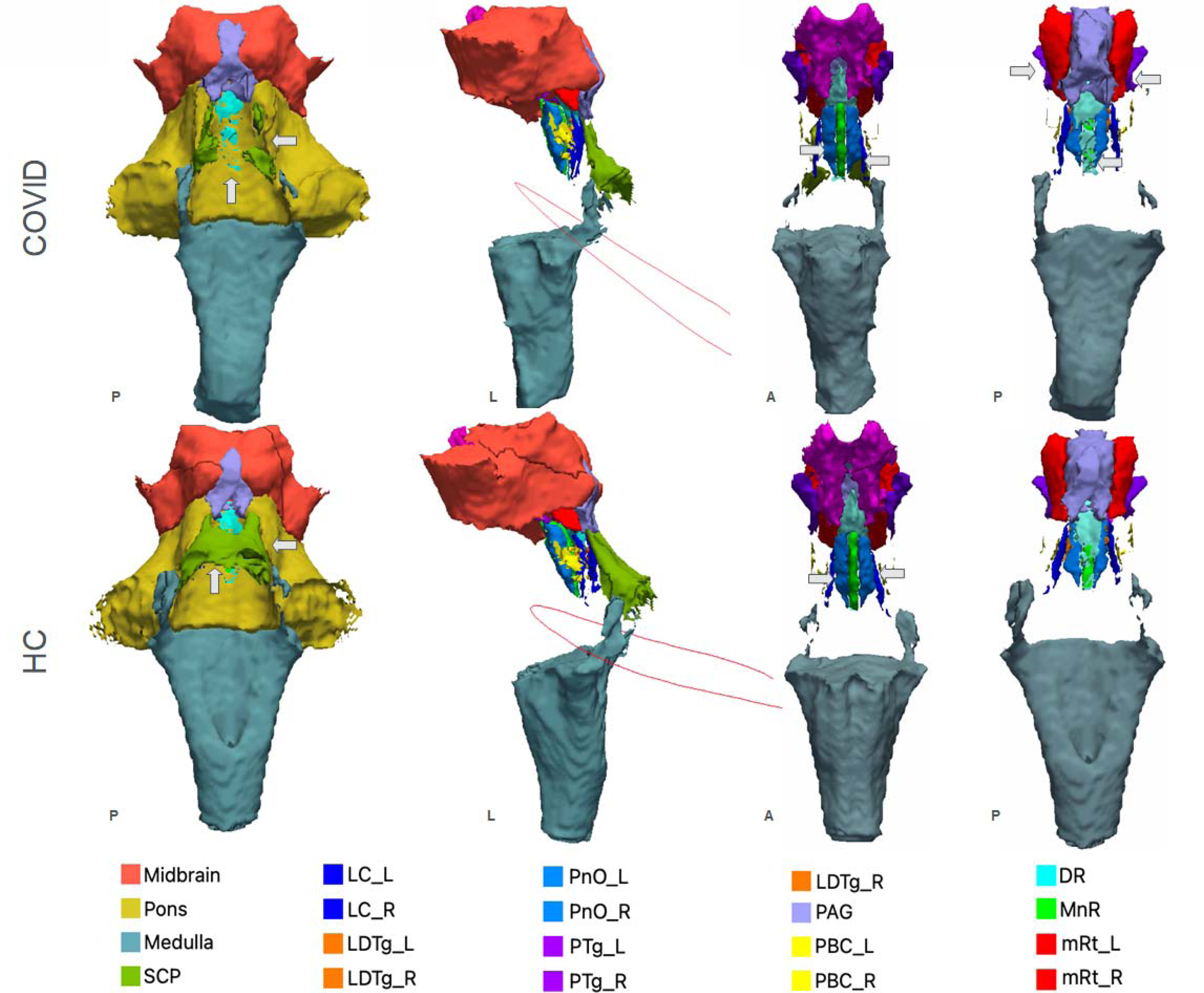
3D Freeview of the structure and volume analysis as isosurface view of the brainstem regions of interest extracted from FreeSurfer segmentation for the COVID group and the healthy control group. The COVID (coronavirus disease 2019) group shows significant volume and structural decrease specifically in the Superior Cerebellar Peduncle (green SCP), Middle Cerebellar Peduncle FA Avg (fractional anisotropy of the conductivity value marked by the red line in the PONS area), dorsal and median Raphe (bright blue/bright green) and Midbrain reticular formation (red)(grey arrows). P = posterior; L = left; A = anterior.

### Volume Reductions and White Matter Integrity Changes

Significant volume loss was observed in the superior cerebellar peduncle (SCP) in LC patients compared to healthy controls (LC: 219.74 mm³ vs. controls: 347.03 mm³, p < .001, Hedges’ g = 3.31). Furthermore, reduced volumes were evident in the dorsal raphe (DR) and midbrain reticular formation (mRt) (p < .001). These changes can be seen in **Figure 3**. In addition to volume changes, decreased fractional anisotropy (FA) was found in the middle cerebellar peduncle (MCP) (LC: 0.39 vs. controls: 0.45, p < .001, Hedges’ g = 1.77), indicating impaired connectivity.

**Figure 3:**
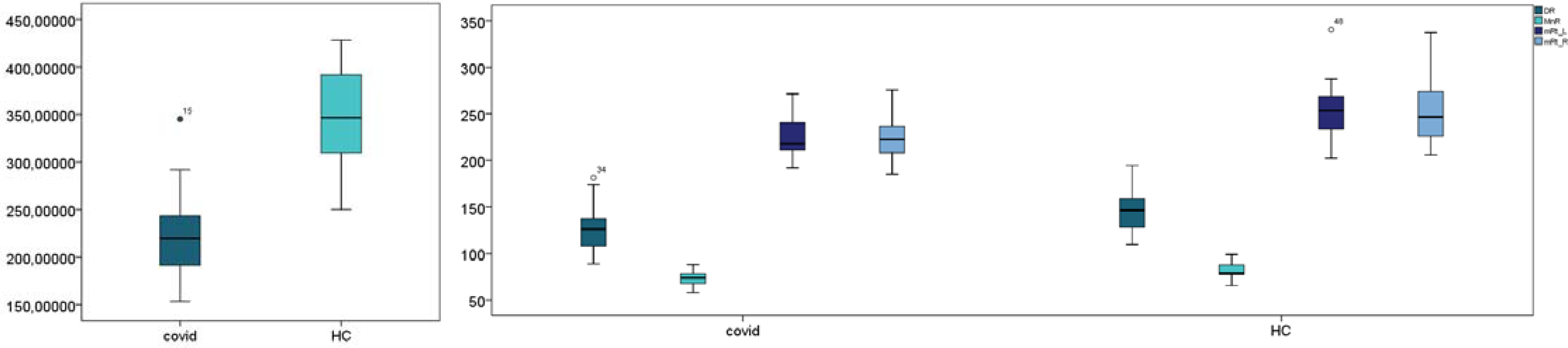
Box plot volume analysis in the region of superior cerebellar peduncle is significant p = <.001 in COVID patients compared to healthy control group. Likewise, in the figure above you can see the missing structure of the upper cerebellar peduncle (grey arrows) shown in green part compared to the structural integrity in the healthy group. Structural and volume changes in the isosurface view can also be seen in the dorsal median raphe and the area of the midbrain reticular formation using 3D imaging. Box plots of the differences of the volume in the dorsal, median Raphe and the Midbrain reticular formation compared to the COVID group and the healthy control group.

### Structural Disruptions and Clinical Correlations

Three-dimensional reconstructions revealed deformities in the 4th ventricle and cerebellar peduncles, suggesting impaired cerebrospinal fluid dynamics as seen in **Figure 4**, which illustrates the structural dissolution and volume reduction of the 4th ventricle in the superior cerebellar peduncle in one and the same COVID patient over time [Iglesias et al., 2015].

**Figure 4:**
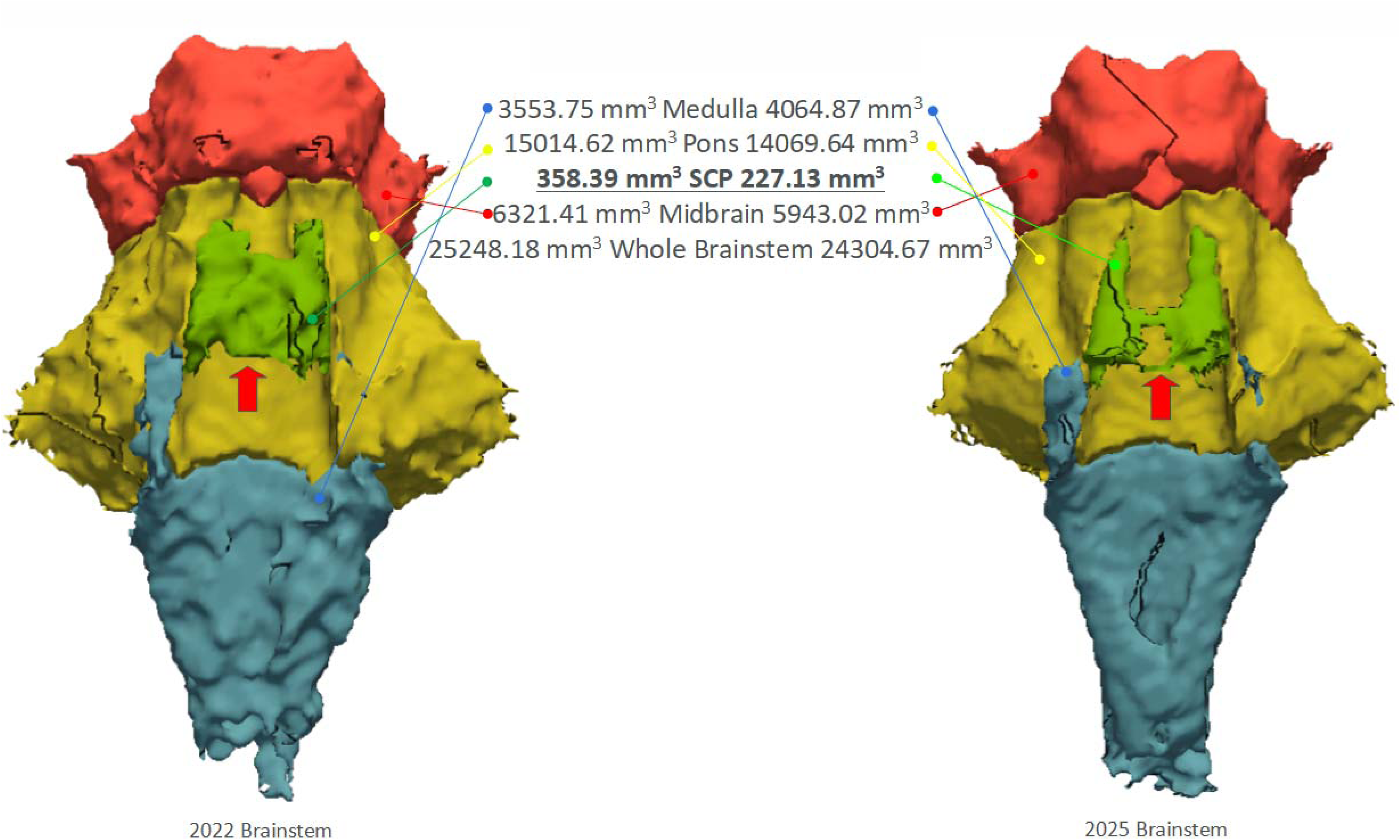
3D Freeview of the structure and volume analysis as isosurface view of the brainstem regions of interest SCP from 2022 to 2025 [Iglesias et al., 2015]. Clearly shows the structural dissolution and volume reduction of the 4th ventricle in the superior cerebellar peduncle in one and the same COVID (coronavirus disease 2019) patient.

Importantly, these volume reductions and FA changes correlated with motor deficits, proprioceptive dysfunction, and autonomic dysregulation.

Scatter plots comparing middle cerebellum peduncle conductivity (FA Avg) with superior cerebellar peduncle volume showed an inverse relationship in the COVID group, suggesting an imbalance in the 4th ventricle (**Figure 5**). The box plots in this figure further highlight the reduction in fractional anisotropy between the brainstem and cerebellum in the COVID group compared to healthy controls [Balsak et al., 2025].

**Figure 5:**
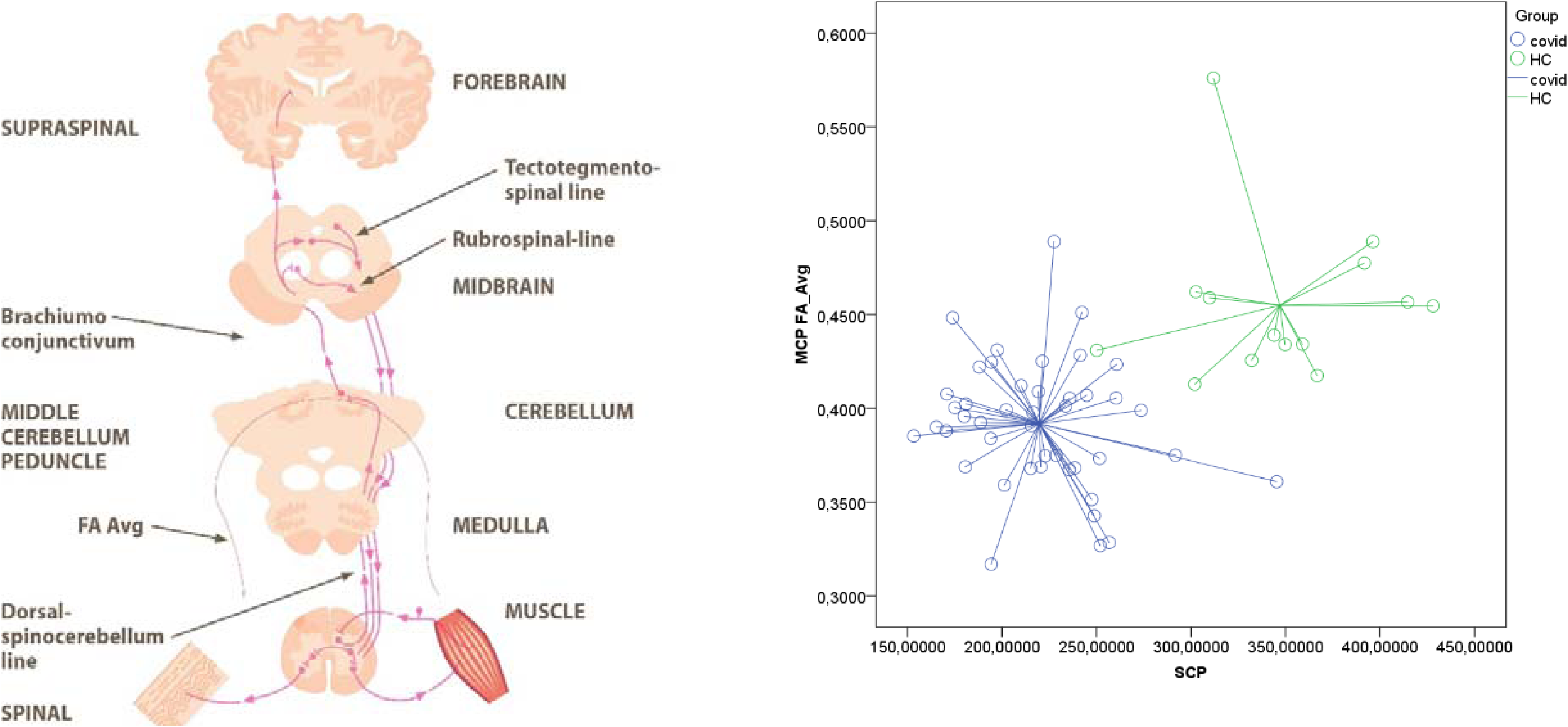
Scatter plots of the average of middle cerebellum peduncle conductivity (FA Avg = fractional anisotropy in their average of conductivity in the vector areas 53 to 65 shown in zentroid) a comparison of the determined volume of the upper cerebellar peduncle of the superior cerebellum peduncle shows that the higher the conductivity in that area in the COVID group (blue points and lines), the lower the volume of the upper cerebellar peduncle and vice versa.

### Implications of the “Broken Bridge Syndrome”

The “Broken Bridge Syndrome” suggests that disruptions in CSF flow dynamics, particularly within the fourth ventricle, may trigger neurodegenerative processes. The presence of autoantibodies and spike protein epitopes could exacerbate CSF stasis, leading to the breakdown of critical neural structures. This syndrome highlights the complex interplay between autoimmune responses, viral protein interactions, and CSF dynamics, offering a new perspective on the neurological sequelae observed in long COVID patients. The gamma-loop shows the pathway of the SARS-CoV-2 virus through the body to the affected areas (**Figure 6**). The brainstem is very important to maintain neural connectivity and homeostasis.

**Figure 6:**
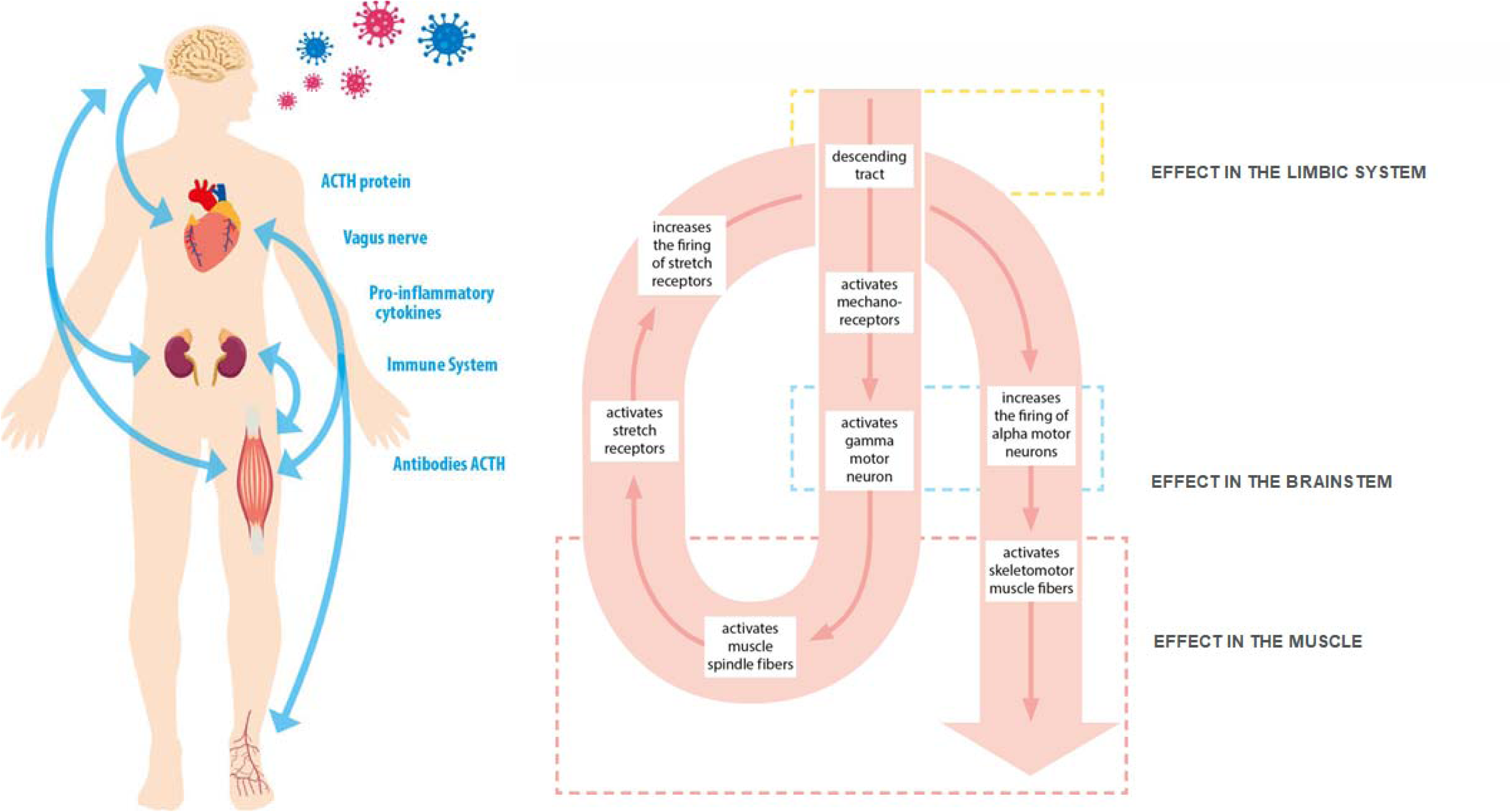
The gamma - loop. the small rectangle at the top represents the limbic system (amygdala); The small rectangle in the center represents the effects that take place in the brainstem or spinal cord, the large rectangle represents the effects in the muscle [Laube et al., 2009]. On the left it shows the path of the SARS-COV2 virus through the body to the places that are affected. 1. Volume Reductions:

- Significant volume loss in the SCP (LC: 219.74 mm³ vs. controls: 347.03 mm³, p < .001).
- Reduced volumes in the dorsal raphe (DR) and midbrain reticular formation (mRt) (p < .001).
2. White Matter Integrity:

- Decreased FA in the MCP (LC: 0.39 vs. controls: 0.45, p < .001), indicating impaired connectivity.
3. Structural Disruptions:

- 3D reconstructions revealed deformities in the 4th ventricle and cerebellar peduncles, suggesting impaired cerebrospinal fluid dynamics.
4. Clinical Correlations:

- Volume reductions and FA changes correlated with motor deficits, proprioceptive dysfunction, and autonomic dysregulation.

In conclusion, these findings underscore the brainstem’s critical role in maintaining neural connectivity and homeostasis. The “Broken Bridge Syndrome” presents a compelling hypothesis linking CSF flow abnormalities, autoimmune responses, and structural degradation, offering a new framework for understanding the neurological complications associated with long COVID and potentially other neurodegenerative conditions. Further research is needed to clarify the mechanisms underlying this syndrome and to explore therapeutic interventions targeting CSF dynamics and autoimmune responses.

## Discussion

Our findings reveal significant structural and functional alterations in the brainstem and cerebellar peduncles of LC patients, supporting the “Broken Bridge Syndrome” hypothesis. The observed volume reductions in the SCP (LC: 219.74 mm³ vs. controls: 347.03 mm³, p < .001, Hedges’ g = 3.31), MCP (LC: 0.39 vs. controls: 0.45, p < .001, Hedges’ g = 1.77) and midbrain reticular formation suggest disrupted motor and autonomic networks, consistent with LC symptoms such as fatigue, motor dysfunction, and sleep disturbances.

### Neuroinflammation and Microglial Activation

These structural changes may be linked to neuroinflammatory processes triggered by SARS-CoV-2 infection. Post-mortem studies have revealed the activation of microglia and infiltration of cytotoxic T lymphocytes predominantly in the brainstem and cerebellum of deceased COVID-19 patients, with limited involvement of the frontal lobe [Matschke et al., 2020]. The virus may enter the blood-brain barrier, initiate microglia activation, and release pro- and anti-inflammatory cytokines, leading to the production of toxic autoantibodies. Radtke et al. (2024) also found activation of intracellular signaling pathways and the release of immune messengers in the brainstem of deceased COVID patients. Although genetic material from deceased COVID patients was detectable, the researchers did not find any infected neurons. Williams et al. (2014) S1 in a virus model shown using the example of the HIV virus blocking antibodies that prevent brain entry. The example, HIV virus can then infect macrophages, microglia, and a small percent of astrocytes (Williams et al. 2014). The activated cells produce cytokines, including TNF-α, IL-1β, IL-6 and IL-8 and chemokines. COVID and its SPIKE proteins use similar mechanisms as we suspect.

### Disrupted Motor and Autonomic Networks

The observed volume loss in the SCP is particularly relevant, given its role in motor coordination and cerebellar communication. Decreased FA in the MCP further supports the notion of impaired white matter integrity, disrupting the flow of information between the cerebellum and brainstem. The volume loss observed in the dorsal raphe (DR) and midbrain reticular formation (mRt) suggests disrupted pain modulation and sleep-wake regulation. The COVID-19 virus may entert the blood-brain barrier by initiates microglia activation and released pro- and anti-inflammatory cytokine storm and leads to toxic autoantibodýs production.

### Clinical Implications and Therapeutic Strategies

The clinical correlations between volume reductions, FA changes, and LC symptoms underscore the functional significance of these structural alterations. Motor deficits, proprioceptive dysfunction, and autonomic dysregulation are common complaints among LC patients, and our findings suggest that the “Broken Bridge Syndrome” may contribute to these symptoms.

Broken Bridge Syndrome testing using FA_Avg fractional anisotropy average is performed using FreeSurfer Tracula, whose segmentation analyzes two regions of the cerebellar peduncle of the 4th ventricle. The middle cerebellar peduncle with FA and the complete brainstem segmentation nuclei of the volume and structure of the rhomboid fossa/cerebellar velum roof of the superior cerebellar peduncle are visualized using isosurface 3D view. DTI segmentation, MPRAG 3Tesla FreeSurfer, and horos data transformation are suitable for this purpose.

The implications of our findings extend beyond LC, as disruptions in CSF flow dynamics and neuroinflammation are implicated in other neurodegenerative conditions [citation needed].

Therapeutic interventions targeting autonomic imbalance, such as vagus nerve stimulation, may offer symptomatic relief by modulating neuroinflammatory responses and improving autonomic function [Stavrakis et al., 2018].

### Limitations and Future Directions

This study has some limitations, including the relatively small sample size and the lack of longitudinal data. Future research should focus on validating these findings in larger cohorts and investigating the long-term effects of SARS-CoV-2 infection on brainstem structure and function. Additionally, further studies are needed to elucidate the precise mechanisms underlying the “Broken Bridge Syndrome” and to identify potential therapeutic targets.

Additionally, laboratory findings correlate with Broken Bridge Syndrome through:

1. Toxic elevated autoantibody AABs (Beta-2 and M-2)
2. Specific epitopes of the COVID virus’s SPIKE protein
3. Cytokine storm of IL-1, IL-6, and IL-8 in their increased numbers (1,000->10,000)

In conclusion, our study provides novel insights into the neuroinflammatory sequelae and structural alterations in the brainstem and cerebellar peduncles of LC patients. The “Broken Bridge Syndrome” hypothesis offers a framework for understanding the complex interplay between viral infection, autoimmune responses, and neurological symptoms. Further research is warranted to develop effective management strategies and improve the quality of life for individuals affected by LC.

## Data Availability

All data produced in the present study are available upon reasonable request to the authors
All data produced in the present work are contained in the manuscript

The incorporation of the attached article enhances our understanding of Post-COVID Syndrome (PCS) and its relationship to the Extended Autonomic System (EAS). This additional information provides valuable insights into the potential mechanisms underlying the long-term effects of COVID-19.

## Introduction

Long COVID, or Post-COVID Syndrome (PCS), has emerged as a significant health concern following the acute phase of SARS-CoV-2 infection. PCS is characterized by persistent symptoms lasting more than 12 weeks post-infection, affecting multiple organ systems and significantly impacting patients’ quality of life1.

Recent research suggests that chronic activation of the Extended Autonomic System (EAS) may play a crucial role in the development and persistence of PCS symptoms. The EAS encompasses not only the traditional components of the autonomic nervous system (sympathetic, parasympathetic, and enteric) but also includes neuroendocrine and neuroimmune systems1.

### The Extended Autonomic System (EAS) in PCS

The EAS concept, as proposed by Goldstein et al., provides a comprehensive framework for understanding the complex interplay between various physiological systems affected in PCS. Key components of the EAS include:

1. Sympathetic adrenergic system
2. Hypothalamic-pituitary-adrenal (HPA) axis
3. Arginine vasopressin system
4. Renin-angiotensin-aldosterone system
5. Cholinergic anti-inflammatory pathway
6. Sympathetic inflammasomal pathway1

These systems are regulated by a hierarchical brain network known as the “central autonomic network”1.

### Pathophysiological Mechanisms

#### HPA Axis Dysregulation

One of the most consistently observed biological abnormalities in both Chronic Fatigue Syndrome (CFS) and PCS is the dysregulation of the HPA axis. Hypocortisolism has been reported in both CFS and COVID-19 populations, suggesting a potential shared mechanism1.

#### Autoantibodies

Recent studies have identified the presence of α- and β-adrenoreceptor and muscarinic receptor autoantibodies in both CFS/ME patients and those recovering from COVID-19. These findings support the hypothesis of autonomic dysfunction and potential autoimmune involvement in PCS1.

#### Viral Immune Evasion

SARS-CoV-2 employs unique strategies to evade the host immune response, including the expression of amino acid sequences that mimic host adrenocorticotropic hormone (ACTH). This mimicry leads to the production of antibodies that inadvertently target the host’s ACTH, potentially contributing to long-term dysregulation of the stress response1.

#### Clinical Manifestations and Prevalence

PCS presents with a wide range of symptoms, including fatigue, dyspnea, chest pain, cognitive impairment, and autonomic dysfunction. Recent studies have reported varying prevalence rates:

- An Italian study found that 53% of patients experienced fatigue, 43% dyspnea, and 22% chest pain at 8 weeks post-infection1.
- A Dutch study reported fatigue in 87.8% of patients, shortness of breath in 74.2%, and chest pressure in 45.4% almost three months after initial symptoms1.

### Proposed Research Directions

To further elucidate the role of EAS activation in PCS, we propose the following research strategies:

1. Longitudinal studies assessing baseline autonomic function, stress levels, and neuroendocrine markers at multiple time points post-infection (3, 6, and 12 months).
2. Investigation of potential prognostic biomarkers related to EAS activation.
3. Evaluation of heart rate variability (HRV) parameters, including:
  - Chronotropic response index
  - Orthostasis test results
  - Analysis of ectopic beats
  - RMSSD and pNN50 values1
4. Assessment of muscle tension and autonomic nervous system regulation ability using specialized techniques developed at the Prof. Stark Institute1.

## Conclusion

The integration of the EAS concept into our understanding of PCS provides a novel framework for investigating the complex interplay between viral infection, immune response, and long-term autonomic dysfunction. By focusing on the chronic activation of the EAS, we may uncover new therapeutic targets and prognostic indicators for PCS patients.

Future research should aim to elucidate the specific mechanisms of EAS dysregulation in PCS and explore potential interventions targeting these systems. This approach may lead to more effective management strategies for the growing population of individuals affected by long-term sequelae of COVID-19.

